# Genome-Wide Polygenic Risk Score Identifies Individuals at Elevated Parkinson’s Disease Risk

**DOI:** 10.1101/2020.10.16.20212944

**Authors:** Yingnan Han, Erin Teeple, Srinivas Shankara, Mahdiar Sadeghi, Cheng Zhu, Dongyu Liu, FinnGen, Clarence Wang, Francesca Frau, Katherine W. Klinger, Stephen L. Madden, Deepak Rajpal, S. Pablo Sardi, Dinesh Kumar

## Abstract

Parkinson’s Disease (PD) is the second most common and fastest-growing neurological disorder. Polygenic Risk Scores (PRS) using hundreds to thousands of PD-associated variants support polygenic heritability. Here, for the first time, we apply a genome-wide polygenic risk score approach using 6.2 million variants to compute a PD genome-wide polygenic risk score (PD-GPRS) via the LDPred algorithm. PD-GPRS validation and testing used Accelerating Medicines Partnership – Parkinson’s Disease (AMP-PD) and FinnGen Consortia genomic data from 1,654 PD Cases and 79,123 Controls. PD odds for the top 8%, 2.5%, and 1% of PD-GPRS were three-, four-, and seven times greater compared with lower percentiles, respectively (p<1e-10). PD age of onset and MDS-UPDRS motor scores also differed by PD-GPRS decile. Enrichment for phagosome related, dopamine signaling, immune related, and neuronal signaling pathways was found for genes nearest high PD-GPRS variants identified by MAF analysis. PD-GPRS offers a promising screening tool to identify high-risk individuals for preventive lifestyle or new drug therapy trials.

**In Brief:** In Han and Teeple et al., Parkinson’s Disease inherited risk is quantified by a genome-wide polygenic risk score (PD-GPRS) approach using 6.2 million variants and data from 80,777 individuals. For the top 2.5% and 1% of PD-GPRS, individuals had five- and seven-fold greater odds of PD, respectively. PD-GPRS was found to be associated with overall PD risk, earlier age of onset, and MDS-UPDRS motor scores. Genes nearest to variants observed at higher frequencies among high-GPRS individuals are enriched for PD-implicated pathways.

**HIGHLIGHTS:**

- Parkinson’s Disease genome-wide polygenic risk score (PD-GPRS) calculated from 6.2 million variants identifies individuals with inherited clinically significant increased neurodegeneration risk.
- Top percentile PD-GPRS individuals were found to have up to seven-fold greater odds of PD and earlier age at PD diagnosis.
- PD-GPRS scores correlated with all-subjects cohort mean MDS-UPDRS motor scores.
- Pathway analysis of genes adjacent to frequently occurring variants in the high PD-GPRS population identified polygenic risk contributions for variations in PD-implicated pathways including dopamine signaling, immune responses, and autophagy pathways.

## INTRODUCTION

Parkinson’s Disease (PD) is the most common neurogenerative disorder after Alzheimer’s Disease and the fastest-growing neurological disorder in the world, with global PD cases doubling to more than 6 million from 1990 to 2015 and a further exponential increase to more than 12 million predicted by 2040 [1-3]. There is currently no objective laboratory or imaging test available to diagnose PD, and early PD symptoms overlap with those of a number of other neurological conditions, making early accurate diagnosis challenging [4]. Treatment guidance and investigations of potential disease-modifying therapies would benefit from tools which improve early diagnosis and enhance profiling of at-risk patients.

Inherited PD susceptibility was first established by the identification of high-penetrance mutations in genes such as SNCA, PARK2, PARK7, PINK1, GBA, and LRRK2, which confer multi-fold increased PD risk, but family history and/or identified mutations occur only in about 5-10% of PD cases [1, 2, 5-8]. Odds of PD vary by mutation severity. For perspective, a meta-analysis of GBA mutation carriers including 11,453 PD patients and 14,565 controls found odds ratios for PD ranging from 2.84-4.94 for mild GBA mutation carriers and from 9.92 - 21.29 for severe GBA mutation carriers [9]. A substantial body of recent work has then focused on understanding the contributions of polygenetic architectures to disease susceptibility. An influential role for polygenetic inheritance in PD is strongly supported by a number of studies which have found significant associations between disease risk, age of onset, motor progression, and cognitive decline with polygenic risk scores (PRS) calculated based on genome-wide association study (GWAS) summary statistics for PD derived from a number of different study populations [10-15].

To date, the largest and most recent genome-wide association study (GWAS) for PD is a 2019 meta-analysis conducted by Nalls et. al. which reports association statistics for 7.8 million single nucleotide polymorphisms in 37,688 PD cases; 18,618 UK Biobank ‘proxy’ cases defined as non-PD cases with a first-degree relative PD case; and 1.4 million control subjects without PD [16]. As part of their meta-analysis, Nalls et. al. computed 88- and 1805-variant PRS models from their summary statistics, including the most highly significant variants in their PRS calculations. AUC performance for PD case identification by these models was found to be 0.65 and 0.69, respectively, corresponding to an estimated genetic liability for PD ranging from 16-36% percent [16].

Previous PRS analyses in PD have utilized up to a few hundred thousand variants for risk score calculations. While these studies have found significant associations between polygenetic features and PD susceptibility [10-12], there remains a need for further analyses which further account for the summary contribution of genomic variation to PD risk. Khera et. al., from Keithersan Lab at the Broad Institute pioneered the application of genome-wide genetic risk analyses for more precise estimation of polygenetic disease risk [17], calculating risk scores from millions of variants, rather than using only subsets of hundreds to thousands of risk alleles [18]. LDPred is one method for such genome-wide polygenic risk estimation and was introduced by Vilhjalmsson and colleagues [19]. This approach infers posterior mean effects for genetic markers using a Gaussian distribution to refine the estimate of the posterior probability of disease while accounting for linkage disequilibrium (LD) [19].

In this analysis, we apply the LDPred algorithm [19] to compute a genome-wide polygenic risk score (GPRS) for PD with the aim of developing a more precise individual-level PD risk estimation tool. For PD-GPRS validation and testing, we used an independent target dataset[20] comprised of 80,777 individuals (1,654 PD Cases: 79,123 Controls) drawn from participants in the Accelerating Medicines Partnership: Parkinson’s Disease (AMP-PD) study [21] and the FinnGen genomics research project [22] whose data had not been used in Nalls et al. PD-GWAS summary statistic calculations. With this analysis, we identified a unique population of patients at substantial and significantly increased risk for Parkinson’s Disease. Cumulative PD incidence plots by PD-GPRS also demonstrate differentiated trajectories, with top and intermediate PD-GPRS percentiles found to have greater incidence by age and earlier disease onset compared with individuals in the lowest scoring decile. Intriguingly, in a further exploratory analysis of AMP-PD cohort data, MDS-UPDRS motor scores were also observed to trend with GPRS score decile, a pattern observed even among mid- and lower-score groups. These findings support the validity of PD-GPRS as a potential tool for more precise and accurate PD risk quantification. Further potential applications for the PD-GPRS are risk stratification for environmental epidemiology studies, clinical screening, and identification of individual patients for yet-to-be identified targeted therapies.

## RESULTS

For this analysis, we used data from the FinnGen (1616 PD Cases and 93,663 Controls) and AMP-PD (1606 PD Cases and 1026 Controls) studies. We considered for inclusion all participants with age ≥ 19 years. Table 1 presents dataset characteristics. However, data from subsets of these studies had been used for GWAS summary statistic calculations [16, 23]. Overlap between GWAS base samples and target subjects may result in biased overestimation of the association between PRS and the condition of interest [20]. To ensure validation set subjects were not included in GWAS summary statistic calculations, we included from AMP-PD only BioFIND study participants and PPMI participants whose identifier codes were not included in a list of GWAS participant identifiers obtained through correspondence with Dr. Nalls. In addition, as a subset of data from the Finnish Parkinson’s Disease Study was used for GWAS calculation [23], we also excluded all FinnGen subjects whose records indicated they were FINRISK study participants. Pooling the filtered data sets, this left 80,777 individuals (1,654 PD Cases: 79,123 Controls) in the target data set used for PD-GPRS validation and testing. QC filtering applied to the 1000 Genomes reference panel and genomic data from participants from the AMP-PD and FinnGen cohorts identified a total of 6,233,374 SNPs eligible for inclusion in the PD-GPRS model.

LDPred computes a posterior mean effect size for each SNP based on Bayesian methods, and the prior distribution of this effect size is assumed as a point-normal mixture distribution. The final input parameter required to perform this estimation is the fraction of causal variants (ρ), which represents an assumption on the number of variants with non-zero effects in the prior Gaussian distribution. In practice, the true value of ρ is unknown, and thus a range of values in the interval (0, 1] are trialed. Selection of ρ for the final score calculation in this analysis was determined as the value which achieved the highest mean area under the curve (AUC) in fivefold cross-validation for prediction of PD case status as a function of GPRS, age, and gender using a logistic regression model.

To generate a dataset to perform this validation, pooled target data set subjects were randomly assigned to be in either the validation or test datasets in a ratio of 1:2. Data from validation set subjects was used to identify the best-performing ρ for GPRS score calculation using fivefold cross-validation. Results for these trials are presented in Supplementary Figure 1. GPRS scores computed for test subjects using the validated ρ value were used for further downstream performance evaluation tasks which then used test set data with this selected ρ. Figure 1 presents a workflow schematic detailing inputs and tuning parameter increments for each step. PD case status prediction was found to be optimized when assuming the maximum possible fraction of causal variants (ρ=1.0), which achieved mean validation set AUC of 0.77 ± 0.06 (± Standard Deviation (SD)).

**Figure 1.**
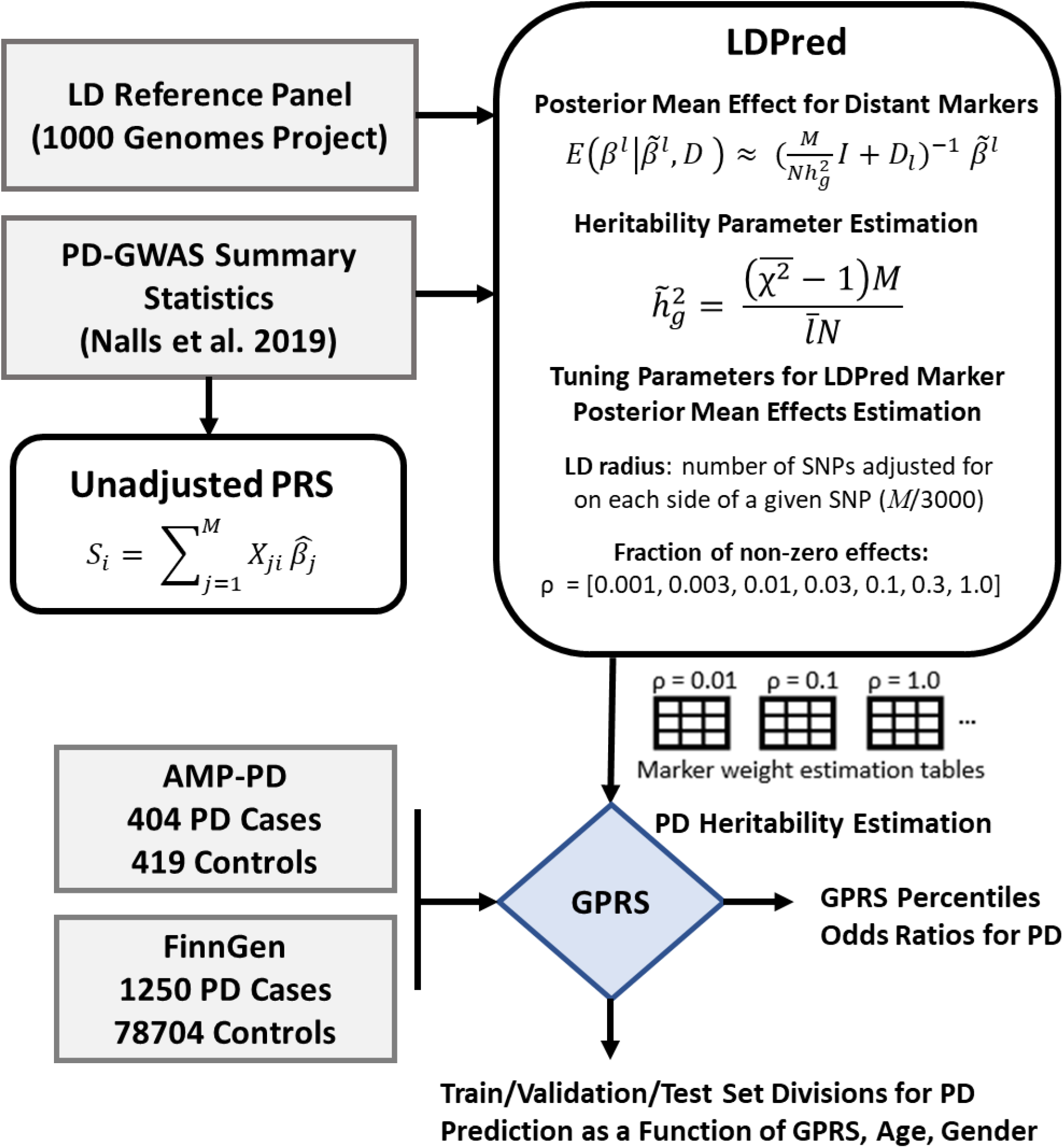
Workflow Schematic for PD-GPRS Validation and Testing. Marker weight estimates are first generated by LDPred for multiple ρ values. GPRS for Case and Control subjects computed based on marker presence/absence and LDPred weights. Odds ratios for PD are reported for top GPRS percentiles. Logistic regression predicting PD case status by GPRS, Age, Gender evaluated by AUC and compared for each ρ.

### PD-GPRS Identifies Individuals at Significantly Increased Risk for Parkinson’s Disease

Examination of PD-GPRS distributions among PD Cases and Controls from the AMP-PD and FinnGen test groups reveals a significant difference in PD-GPRS among PD Cases versus Control in the testing data fraction (Fig. 2A; p <1e-10). Prevalence of PD was also found to increase sharply among individuals in the highest PD-GPRS percentiles (Fig. 2B). In comparisons between each of the top percentiles and their lower percentile complements, odds ratios for PD were substantially and significantly greater for the high-percentile PD-GPRS groups, with subjects in the top 8%, 4.5%, 2.5% and 1% of normalized PD-GPRS found to have 3.1, 4.1, 5.2, and 7.9 times significantly greater odds of PD, respectively (Fig. 2C; p<1e-10).

**Figure 2.**
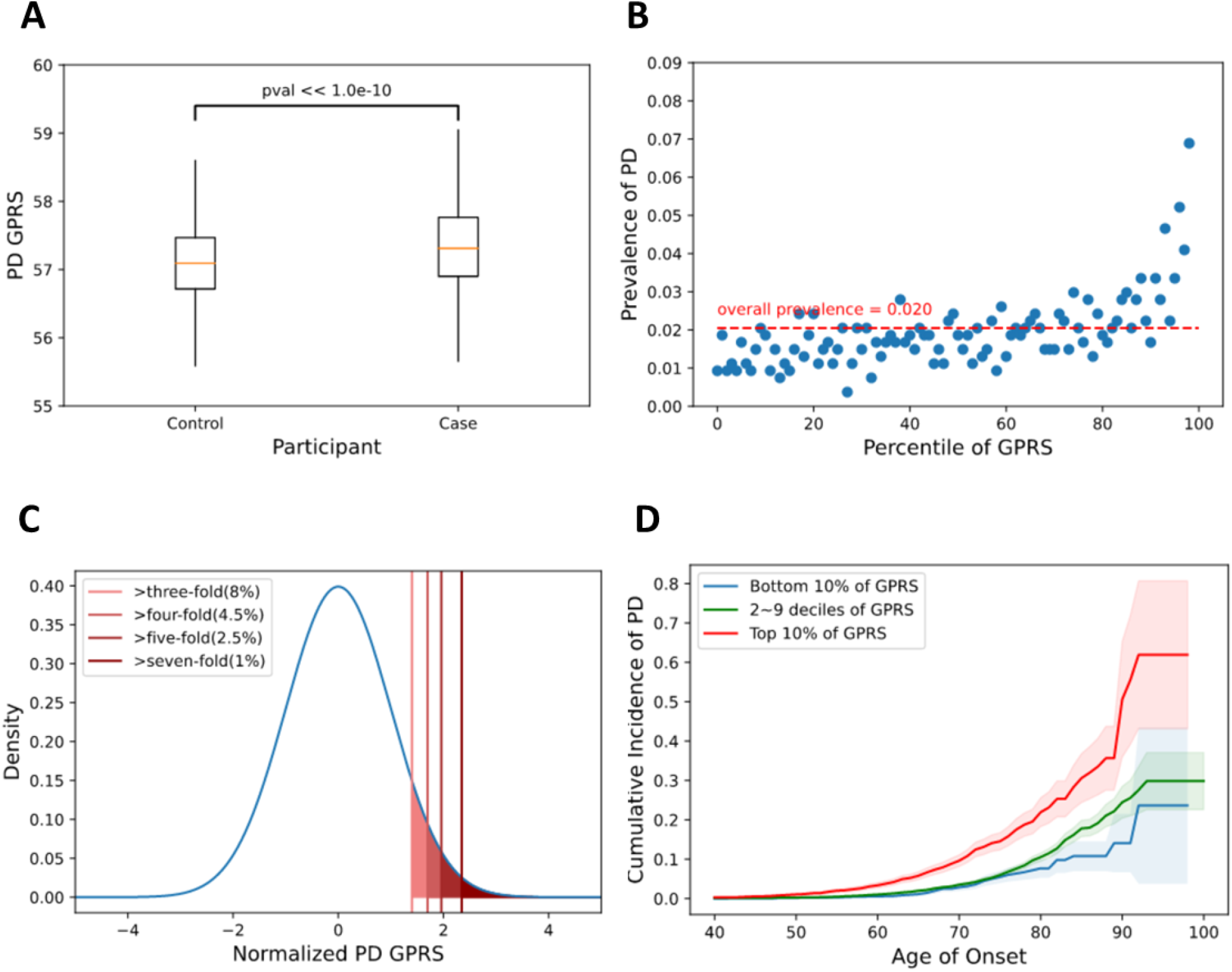
Individuals with high GPRS are at greater risk of PD. GPRS score distributions differ significantly between PD Cases and Controls with higher mean GPRS in PD cases (A). Prevalence of PD increases with increasing GPRS percentile (B). Individuals in the top 8%, 4.5%, 2.5%, 1% of GPRS have 3, 4, 5, 7-fold greater odds of PD versus the rest of the study cohort (C) Cumulative incidence of PD by age of onset and GPRS percentile shows earlier onset age and greater prevalence with higher GPRS (D).

### PD-GPRS Is Associated with Disease Prevalence, Age of Onset, and MDS-UPDRS Motor Scores

As data from these pooled study cohorts was cross-sectional, cumulative incidence plots were constructed for PD cases using age of onset data. PD-GPRS percentile was also found to be associated with earlier age of disease onset, with higher-score individuals having earlier age of onset and greater cumulative incidence of PD over time (Fig. 2D). Notably, differentiation for PD trajectories in these cumulative incidence plots occurs not only between highest- and lowest-scoring PD-GPRS percentiles, but also between the high, low, and intermediate scoring-groups.

The Movement Disorders Society update of the Unified Parkinson’s Disease Rating Scale (MDS-UPDRS) is among standardized measurements collected for both PD Case and Control participants enrolled in the AMP-PD cohorts. MDS-UPDRS consists of four sections: Part I: non-motor experiences of daily living; Part II: motor experiences of daily living; Part III: motor examination; and part IV: motor complications, with items in each section scored with a rating from 0 (normal) to 4 (severe) [24]. Apart from a given participant’s classification as a PD case versus Control, MDS-UPDRS provides a supplementary assessment of subjective and objective neurological function. Intriguingly, among all AMP-PD subjects, including both PD Cases and Controls in a plot of MDS-UPDRS versus GPRS decile, we observe a graded difference in average MDS-UPDRS scores for parts II and III across PD-GPRS deciles among AMP-PD test group subjects (Fig. 3).

**Figure 3.**
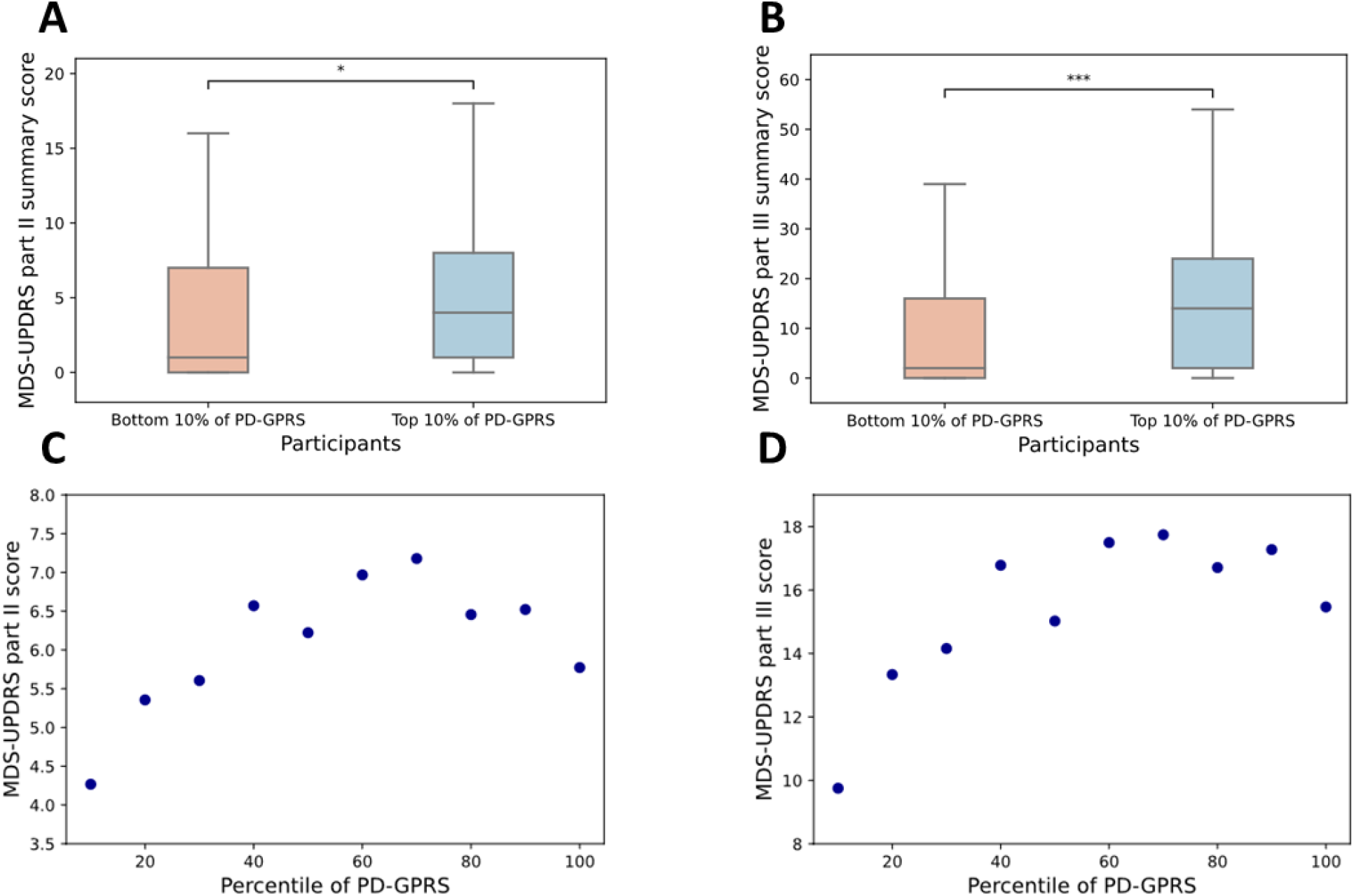
MDS-UPDRS scores among AMP-PD Cases and Controls by GPRS. Permutation testing finds significant differences in clinical scores between High/Low GPRS populations for MDS-UPDRS Parts II and III. Score distributions for MDS-UPDRS Part II (A) and Part III (B) for high and low GPRS. Mean MDS-UPDRS plotted by decile shows a trend for increasing mean Part II (C) and III (D) scores with increasing GPRS.

### Nearest Genes to High PD-GPRS Variants are Enriched for PD-GWAS Loci and PD-Implicated Pathways

In this analysis, we observed significantly increased odds of PD among high-PD-GPRS individuals. Based on this finding, we proposed that alleles observed with higher frequency in high-GPRS populations are those most likely to be related to genes with causal contributions to PD pathogenesis. MAF analyses was then used to identify alleles with the greatest odds of being present in the genomes of individuals in the highest versus lowest 5% of PD-GPRS scores (Fig.4A). Pathway and gene ontology analysis was then performed for the set of the top 1000 genes identified as nearest genes to variants found to occur with significantly greater frequency among high-GPRS individuals in contrast to the low-GPRS cohort. Enrichment network analysis using NetworkAnalyst [25] to profile these genes revealed enrichment of this gene set for a number of PD-implicated pathways, including phagosome, dopaminergic synapse, and GABAergic signaling pathways, as well as peripheral adaptive immunity pathways involved in inflammation (Fig. 4B).

**Figure 4.**
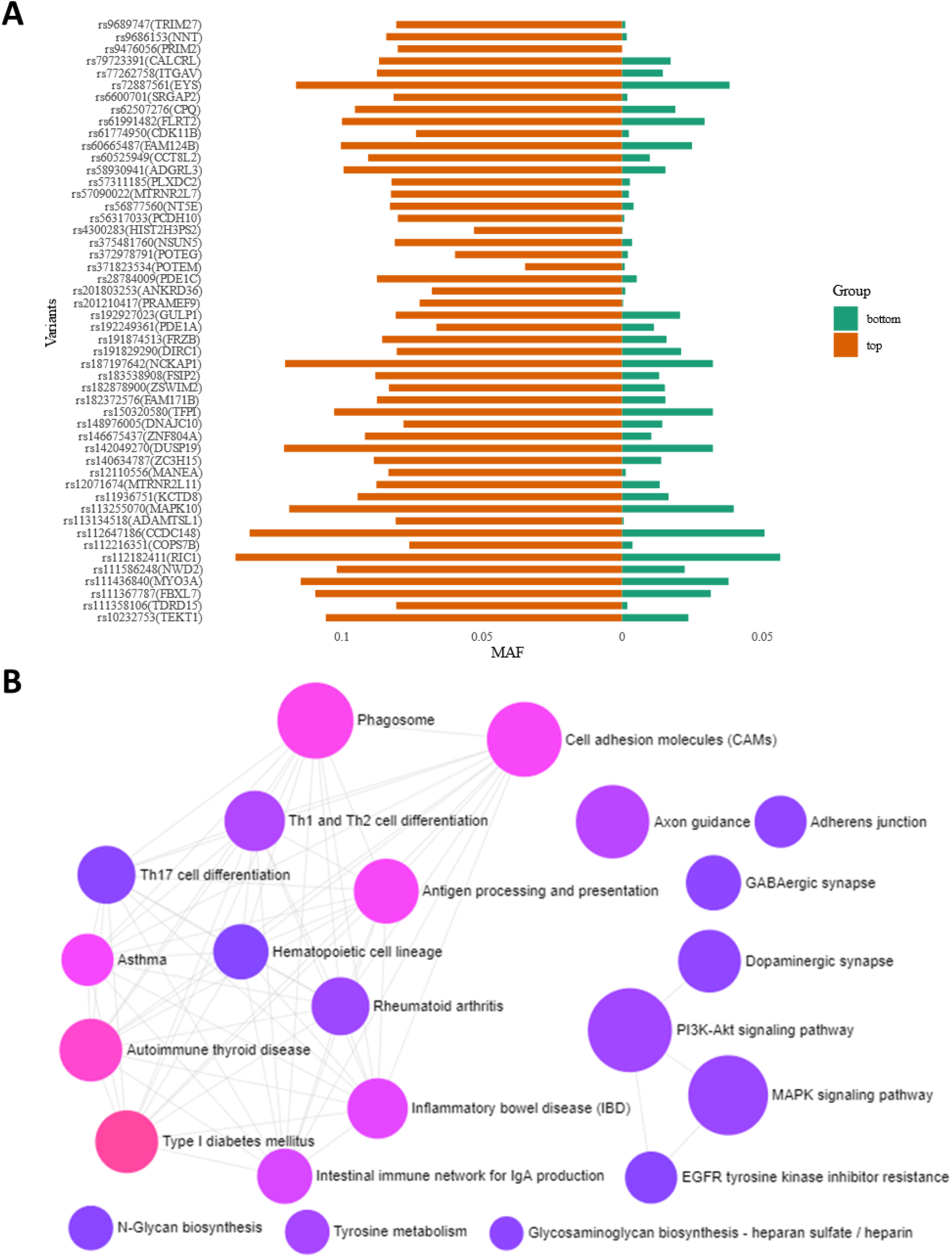
Two-tail analysis comparing variants observed for high versus low PD-GPRS populations from AMP-PD and FinnGen. Top variants identified by MAF as more frequent among high GPRS individuals (A). Canonical pathways identified from top 1000 genes associated with most significant variants are enriched for pathways including cell adhesion, growth, dopaminergic signaling, and immune responses; node size and color saturation indicate greater enrichment statistical significance (cancer and infectious disease pathways excluded) (B).

## DISCUSSION

In this analysis, we describe a new polygenetic PD risk estimation tool which computes PD-GPRS using genomic data for 6.2 million variants using the LDPred method. Subjects in the top percentiles of PD-GPRS were found to have substantially and significantly increased risk for PD, with more than five-fold greater odds of PD in the top 2.5% of scores and seven-fold greater odds of PD in the top 1%. Further, cumulative incidence plots by PD-GPRS decile demonstrated differences in incidence and age of onset trajectories between high, low, and middle PD-GPRS score percentiles, and MDS-UPDRS motor scores were found to trend with PD-GPRS, as well. These results provide strong evidence for the influence of complex inheritance in determining PD susceptibility and invite further questions regarding whether heterogeneous PD polygenetic subtypes may exist.

Estimated genetic heritability for PD calculated by LDPred in our study is 47.6%. This estimate is higher than the PD genetic liability estimate of 16-36% reported by Nalls et al. [16] in the study from which we obtained summary statistics used for our PD-GPRS calculations. Although in this study we used the same GWAS summary statistics, the LDPred approach supports the inclusion of a much higher number of variants for polygenic risk calculation, which likely accounts for our greater heritability estimate. Nalls et. al. also evaluated PD case prediction by PRS models using subsets of highly significant variants, achieving an average AUC performance of 0.65 and 0.69 for 88- and 1805-variant PRS, respectively. Including a greater number of variants in our PD-GPRS model (6.2 million) enables us to achieve a slightly higher average test set AUC of 0.77. This finding is consistent with our PD-GPRS model capturing an increased proportion of disease variability attributable to genetic variation. LDPred applications in other disorders have reported similar improvements in GPRS prediction accuracy [19]. Our heritability estimate is also consistent with an upper range estimate for PD heritability reported in another recently published study: in a genome-wide complex trait analysis performed by Keller et al. for a number of different PD population data sets, estimates of PD heritability ranged from approximately 16% in a UK cohort to nearly 49% in a cohort from Finland [26].

Genetic variants associated with greater PD risk have been linked with diverse cellular pathways, including lysosomal storage, lipid metabolism, ion channel function, and oxidative phosphorylation [1, 5, 8]. How individual genetic architectures may contribute to the pathogenesis of PD remains incompletely understood. Several intriguing pathways were found to be enriched for among genes linked to higher frequency PD alleles among top versus bottom percentile PD-GPRS cohorts by MAF analysis. These pathways include pathways related to cell adhesion, Dopaminergic synapses, antigen presentation pathways, neuronal growth and development, and immune responses. Interestingly, genes linked with high PD-GPRS alleles were found to be enriched for pathways linked with multiple autoimmune inflammatory disorders, including inflammatory bowel disease, type I diabetes and autoimmune thyroid conditions. Previous studies have reported associations between PD and inflammatory bowel disease[27] as well as autoimmune thyroid disorders [28]. Another recent study of pleiotropic genetic risk factors identified 17 loci which are shared risk factors for Parkinson disease and type 1 diabetes, Crohn’s disease, ulcerative colitis, rheumatoid arthritis, celiac disease, psoriasis, and multiple sclerosis [29]. In addition, cellular adhesion molecules, which was found to be among the pathways enriched for among the identified genes, has also attracted recent attention for a possible significance in PD [30], further highlighting the novelty and importance of pathways identified through polygenetic risk stratification by PD-GPRS.

Excitingly, as shown in this analysis, PD-GPRS performance for case identification and decile-stratified risk profiling may provide further opportunities to identify and examine the metabolic profiles of individuals with scores which reflect the co-occurrence of varying numbers of alleles with intermediate disease associations in population studies. It is possible that PD genetic risk architectures may harbor heterogeneous subgroups defined by such co-occurrences and that such subgroups might benefit from targeted therapies yet to be developed. Identification and characterization of preclinical metabolic profiles among individuals at varying risk would be beneficial for understanding the interaction of genetic and metabolic conditions which may precede disease onset or identify pathophysiological subtypes. In addition, information obtained from orthogonal sources, such as wearable devices for objective functional motor performance assessment and clinical and family history may be integrated with PD-GPRS for further refinement in diagnostic or screening applications. Such applications would be of immediate use for individuals presenting with undifferentiated neurological syndromes which may be prodromal symptoms of PD or other disorders. Further studies are needed to fully interrogate these possibilities, as well as to replicate these analyses in diverse populations.

## Supporting information

EQUATOR-STROBE checklist

## Data Availability

Analysis code will be made available as supplemental files at the time of publication.
FinnGen data is restricted and requires authorization for access:
https://www.finngen.fi/en/about.
Unified Amp-PD Cohorts data is restricted and requires authorization for access:
https://amp-pd.org/
Information and access to the Terra computing platform can be found at
https://terra.bio/. Terra is developed by the Broad Institute of MIT and Harvard in
collaboration with Verily Life Sciences. The Terra system was used for access to AMP-PD
data and for computing PD-GPRS scores using LDPred algorithm.
LDpred is a Python-based software package which adjusts GWAS summary statistics for
the effects of linkage disequilibrium (LD). The method reference is Vilhjalmsson et al.
(AJHG 2015) [http://www.cell.com/ajhg/abstract/S0002-9297(15)00365-1]. LDPred code
is available at https://github.com/bvilhjal/ldpred.
Parkinsons Disease Genome-Wide Association Study (GWAS) summary statistics from
Nalls et al. used for PD-GPRS calculation using LDPRed are available from a
supplementary table to the publication[16]. Complete summary statistics table:
https://www.biorxiv.org/content/10.1101/388165v3.full.pdf, link to data table file is
found under heading DATA ACCESS.
Genome reference panel source is 1000 Genomes Project phase 3:
https://www.internationalgenome.org/data/.

https://www.finngen.fi/en/about

## ACKNOWLEDGMENTS

Data used in the preparation of this article were obtained from FinnGen Release 4 and the AMP PD Knowledge Platform.

FinnGen: The FinnGen project is funded by two grants from Business Finland (HUS 4685/31/2016 and UH 4386/31/2016) and eleven industry partners (AbbVie Inc, AstraZeneca UK Ltd, Biogen MA Inc, Celgene Corporation, Celgene International II Sàrl, Genentech Inc, Merck Sharp & Dohme Corp, Pfizer Inc., GlaxoSmithKline, Sanofi, Maze Therapeutics Inc., Janssen Biotech Inc). Following biobanks are acknowledged for collecting the FinnGen project samples: Auria Biobank (www.auria.fi/biopankki), THL Biobank (www.thl.fi/biobank), Helsinki Biobank (www.helsinginbiopankki.fi), Biobank Borealis of Northern Finland (https://www.ppshp.fi/Tutkimus-ja-opetus/Biopankki/Pages/Biobank-Borealis-briefly-in-English.aspx), Finnish Clinical Biobank Tampere (www.tays.fi/en-US/Research_and_development/Finnish_Clinical_Biobank_Tampere), Biobank of Eastern Finland (www.ita-suomenbiopankki.fi/en), Central Finland Biobank (www.ksshp.fi/fi-FI/Potilaalle/Biopankki), Finnish Red Cross Blood Service Biobank (www.veripalvelu.fi/verenluovutus/biopankkitoiminta) and Terveystalo Biobank(www.terveystalo.com/fi/Yritystietoa/Terveystalo-Biopankki/Biopankki/). All Finnish Biobanks are members of BBMRI.fi infrastructure (www.bbmri.fi).

AMP PD: Data used in the preparation of this article were obtained from the AMP PD Knowledge Platform. For up-to-date information on the study, https://www.amp-pd.org. “AMP PD – a public-private partnership – is managed by the FNIH and funded by Celgene, GSK, the Michael J. Fox Foundation for Parkinson’s Research, the National Institute of Neurological Disorders and Stroke, Pfizer, Sanofi, and Verily.

AMP PD Cohort: Clinical data and biosamples used in preparation of this article were obtained from the Fox Investigation for New Discovery of Biomarkers (BioFIND), the Harvard Biomarker Study (HBS), the Parkinson’s Progression Markers Initiative (PPMI), and the Parkinson’s Disease Biomarkers Program (PDBP).

BioFIND is sponsored by The Michael J. Fox Foundation for Parkinson’s Research (MJFF) with support from the National Institute for Neurological Disorders and Stroke (NINDS). The BioFIND Investigators have not participated in reviewing the data analysis or content of the manuscript. For up-to-date information on the study, visit michaeljfox.org/biofind.

The Harvard NeuroDiscovery Biomarker Study (HBS) is a collaboration of HBS investigators [full list of HBS investigator found at https://www.bwhparkinsoncenter.org/biobank/] and funded through philanthropy and NIH and Non-NIH funding sources. The HBS Investigators have not participated in reviewing the data analysis or content of the manuscript.

PPMI – a public-private partnership – is funded by the Michael J. Fox Foundation for Parkinson’s Research and funding partners, including [list the full names of all of the PPMI funding partners found at www.ppmi-info.org/fundingpartners]. The PPMI Investigators have not participated in reviewing the data analysis or content of the manuscript. For up-todate information on the study, visit www.ppmi-info.org.

Parkinson’s Disease Biomarker Program (PDBP) consortium is supported by the National Institute of Neurological Disorders and Stroke (NINDS) at the National Institutes of Health. A full list of PDBP investigators can be found at https://pdbp.ninds.nih.gov/policy. The PDBP Investigators have not participated in reviewing the data analysis or content of the manuscript.

The authors thank Dr. Eline Appelmans for her help and support on this project. The authors thank Dr. Mike Nalls and Prof. Andrew Singleton for their help and communications which allowed independent target population identification.

---

**Supplement Figure 1.**
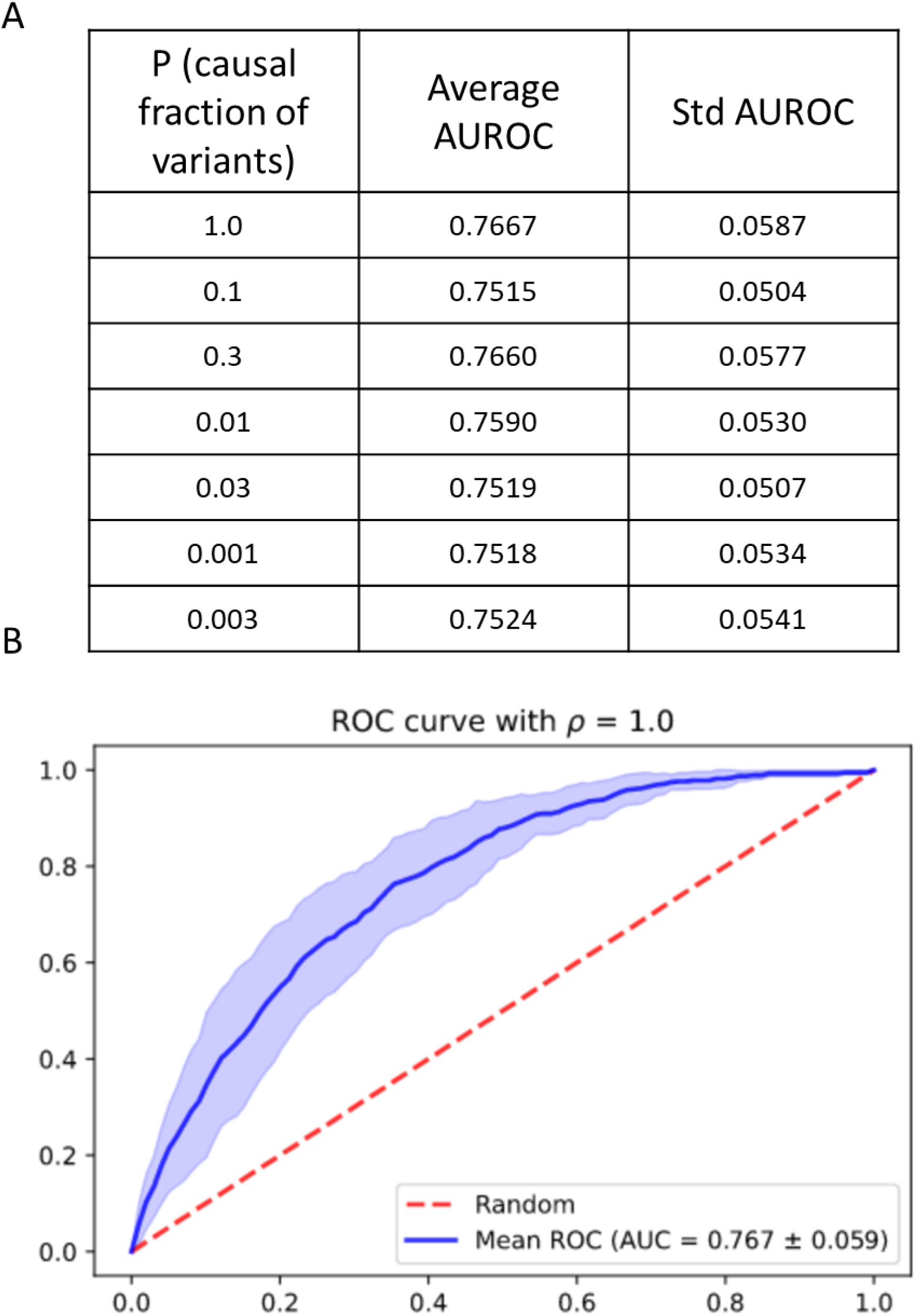
ROC curve in validation dataset. (A) Average AUROC in 5-fold cross validation with different parameters (ρ = 0.001, 0.003, 0.01, 0.03, 0.1, 0.3, 1.0). (B) The ROC curve derived with ρ = 1.0 in 5-fold cross validation.

**Supplement Figure 2.**
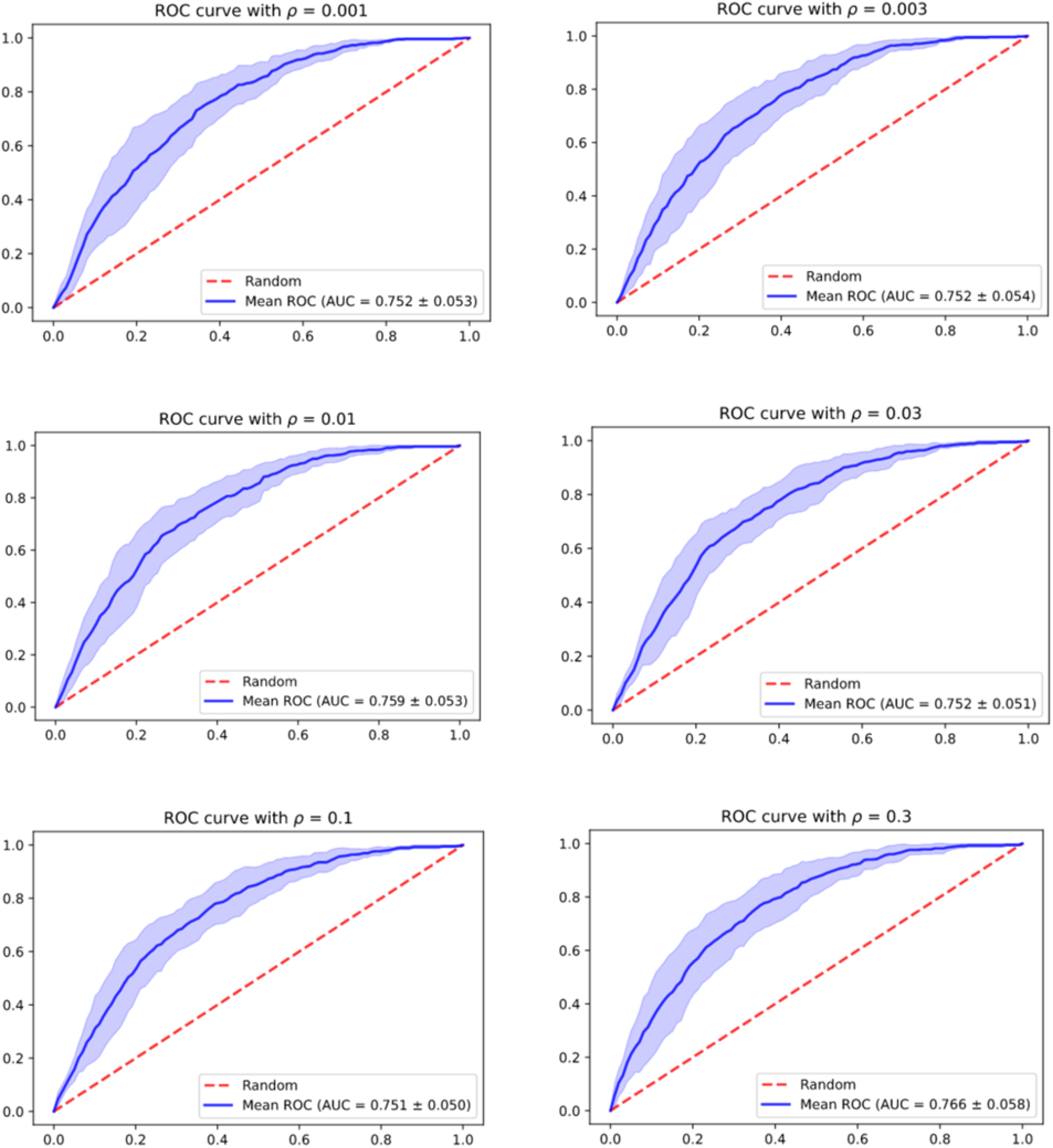
**ROC curve in validation dataset with different parameters** (ρ = 0.001, 0.003, 0.01, 0.03, 0.1, 0.3, 1.0)

**Supplement Table 1.**
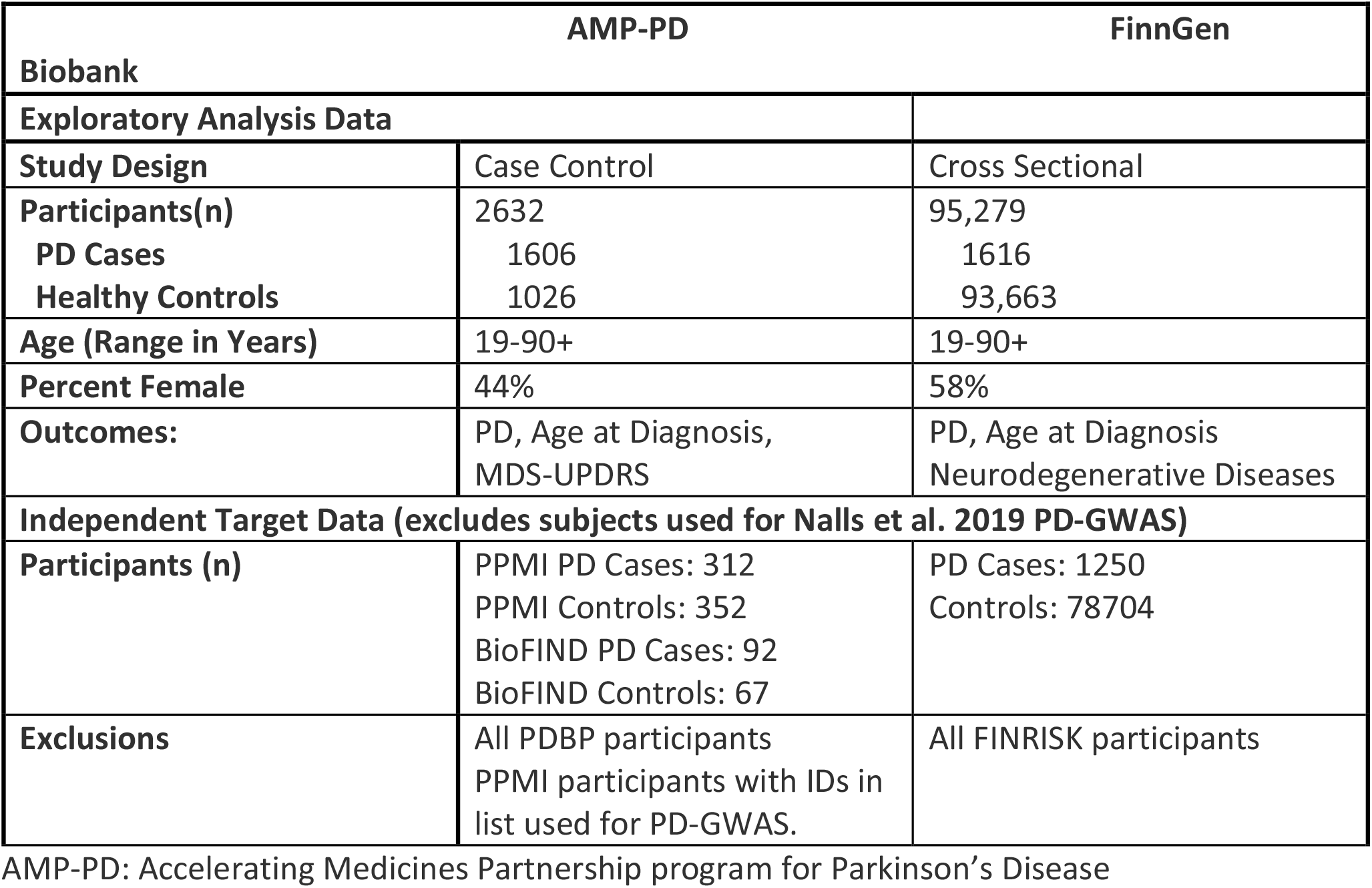
Dataset Characteristics.

## METHODS

### RESOURCE AVAILABILITY

#### Lead Contact

Further information and requests for resources and code files should be directed to the Lead Contact: Dinesh Kumar

#### Materials Availability

- Information and access to the Terra computing platform can be found at https://terra.bio/. Terra is developed by the Broad Institute of MIT and Harvard in collaboration with Verily Life Sciences. The Terra system was used for access to AMP-PD data and for computing PD-GPRS scores using LDPred algorithm.
- LDpred is a Python-based software package which adjusts GWAS summary statistics for the effects of linkage disequilibrium (LD). The method reference is Vilhjalmsson et al. (AJHG 2015) [http://www.cell.com/ajhg/abstract/S0002-9297(15)00365-1]. LDPred code is available at https://github.com/bvilhjal/ldpred.
- Parkinson’s Disease Genome-Wide Association Study (GWAS) summary statistics from Nalls et al. used for PD-GPRS calculation using LDPRed are available from a supplementary table to the publication[16]. Complete summary statistics table: https://www.biorxiv.org/content/10.1101/388165v3.full.pdf, link to data table file is found under heading “DATA ACCESS”.
- Genome reference panel source is 1000 Genomes Project phase 3: https://www.internationalgenome.org/data/.

#### Data and Code Availability

- Analysis code will be made available as supplemental files at the time of publication.
- FinnGen data is restricted and requires authorization for access: https://www.finngen.fi/en/about.
- Unified Amp-PD Cohorts data is restricted and requires authorization for access: https://amp-pd.org/

### METHOD DETAILS

#### Data Sources

Whole-genome sequencing data from PD Case and Control subjects was obtained from participants in two large cohorts, the Accelerating Medicines Partnership: Parkinson’s Disease (AMP-PD) Study [21] and the FinnGen Biobank Cohort [22].

#### The Accelerating Medicines Partnership: Parkinson’s Disease (AMP-PD) Study

AMP-PD is a public-private partnership between the United States National Institutes of Health, non-profit organizations, and pharmaceutical and life sciences companies which aims to identify and validate new, impactful biological targets for PD therapeutics. Using the Terra platform for data access and analysis, AMP-PD makes available unified data from a number of published PD cohort studies: the Michael J. Fox Foundation (MJFF) and National Institutes of Neurological Disorders and Stroke (NINDS) BioFIND study, Harvard Biomarkers Study (HBS), the NINDS Parkinson’s Disease Biomarkers Program (PDBP), and MJFF Parkinson’s Progression Marker Initiative (PPMI) [21].

#### FinnGen

FinnGen is a public-private partnership between the Finnish National Institute for Health and Welfare, universities, biobanks, and a number of pharmaceutical companies which was launched in 2017 with the aim of collecting health and genomic sequencing data from 500,000 individuals for use in analysis to identify new diagnostic methods and therapies [22]. Patients and control subjects in FinnGen provided informed consent for biobank research, based on the Finnish Biobank Act. Alternatively, separate research cohorts, collected prior the start of FinnGen (August 2017), were collected based on study-specific consents and later transferred to the Finnish biobanks after approval by Fimea, the National Supervisory Authority for Welfare and Health. Recruitment protocols followed the biobank protocols approved by Fimea. The Coordinating Ethics Committee of the Hospital District of Helsinki and Uusimaa (HUS) approved the FinnGen study protocol Nr HUS/990/2017.

The FinnGen project is approved by Finnish Institute for Health and Welfare (THL), approval number THL/2031/6.02.00/2017, amendments THL/1101/5.05.00/2017, THL/341/6.02.00/2018, THL/2222/6.02.00/2018, THL/283/6.02.00/2019), Digital and population data service agency VRK43431/2017-3, VRK/6909/2018-3, the Social Insurance Institution (KELA) KELA 58/522/2017, KELA 131/522/2018, KELA 70/522/2019 and Statistics Finland TK-53-1041-17.

The Biobank Access Decisions for FinnGen samples and data utilized in FinnGen Data Freeze 4 include: THL Biobank BB2017_55, BB2017_111, BB2018_19, BB_2018_34, BB_2018_67,BB2018_71, BB2019_7 Finnish Red Cross Blood Service Biobank 7.12.2017, Helsinki Biobank HUS/359/2017, Auria Biobank AB17-5154, Biobank Borealis of Northern Finland_2017_1013, Biobank of Eastern Finland 1186/2018, Finnish Clinical Biobank Tampere MH0004, Central Finland Biobank 1-2017, and Terveystalo Biobank STB 2018001.

Based on our use of a European reference genome panel, for this analysis, eligible subjects were AMP-PD Caucasian/White participants and all FinnGen participants for whom genome sequencing data was available and who were over 19 years of age. A small number of AMP-PD participants with PD prodromal symptoms without a confirmed diagnosis of PD and participants with a PD diagnosis whose diagnosis was changed to another neurological disorder during the study were identified and excluded (n = 79). Eligible participants with a diagnosis of Parkinson’s Disease were classified as PD Cases (AMP-PD Cases: n = 1606; FinnGen Cases: n = 1616) and participants without a Parkinson’s Disease Diagnosis were classified as Controls (AMP-PD Controls: n = 1026; FinnGen Controls: n = 93,663). Subjects whose data was used for PD-GWAS summary statistic calculations were identified by their cohort and/or subject ID (AMP-PD/PPMI participants) (n = 1809) or in the case of FinnGen, indication of FINRISK participation in the subject record (n = 15,325). Excluding these subjects, the independent target dataset used for PD-GPRS validation and testing was comprised of 80,777 individuals (1,654 PD Cases: 79,123 Controls).

#### LDPred PD-GPRS Calculation

GPRS is derived to predict the likelihood of a specific outcome according to cumulative effect of multiple variants. This score is obtained as the aggregation of weighted variants, where the weight represents the strength of variants association with disease risk, which can be derived from a genome-wide association study. PD-GPRS was calculated in this analysis using the LDpred algorithm [19] to generate posterior effect sizes for each SNP. The algorithm computes a posterior mean effect size based on Bayesian methods, with the prior distribution of effect size assumed as a point-normal mixture distribution. In contrast to unadjusted, clumped, and LD-pruned PRS approaches [31, 32], LDPred was proposed as a method by which to reduce loss of information that can occur when variants are thresholded by significance level or relative locations and was explicitly designed to support inclusion of a much higher number of variants for genome-wide polygenic risk score (GPRS) calculation. LDPred has been shown to increase the proportion of variability in disease incidence explained by genomic variation in comparison to unadjusted and LD-pruned PRS, and has been successfully applied for identification of individuals with clinically significant increased risk for diseases including obesity, coronary artery disease, atrial fibrillation, type 2 diabetes, and schizophrenia [17-19].

To determine the prior distribution, LDpred algorithm requires several inputs: PD-GWAS summary statistics for markers to be used for GPRS calculation, a genomic reference panel used to account for linkage disequilibrium, and an estimate (manually adjustable) of the fraction of causal variants, which represents the number of variants with non-zero effects in the prior Gaussian distribution (ρ) [19]. We made experiments with seven different values for the fraction of causal variants, then selected the parameter with best prediction performance in the validation dataset. For this study, we used GWAS summary statistics reported by Nalls et al. for 7.8 million single nucleotide polymorphisms [16]. LDPred also requires input of a reference genome panel [19]. The reference panel used for this analysis consisted of 503 samples from individuals of European ancestry obtained from the 1000 Genomes Project [33]. Reference panel data is used to filter SNPs with high missing rates (>5%) from inclusion in the GPRS calculation. In this preprocessing step, SNPs with low minor allele frequency (MAF) (<0.01) are also filtered, for both GWAS hits and reference panel samples, and ambiguous nucleotides are removed.

Summary statistics reported by Nalls et al. were used as input for the LDPred algorithm in order to estimate variant posterior mean effects. Nalls et al. report summary statistics calculated from 7.8 million single nucleotide polymorphisms in 37,688 cases; 18,618 UK Biobank ‘proxy’ cases defined as non-PD cases with a first-degree relative PD case; and 1.4 million controls. The genomic LD reference panel used for this analysis was a panel of 503 Europeans samples from 1000 Genomes Project [33]. In the reference panel, we filtered out SNPs with high missing rates, which we defined as greater than 5%. We also filtered out SNPs with low MAF, defined as less than 0.01, in either GWAS hits or reference panel. All ambiguous nucleotides (A/T, C/G) were removed, as well.

Case and Control subjects from the two cohorts were pooled and randomized in a 1:2 ratio to either Validation or Testing sets. Validation Set data was used to determine the optimal value for ρ, which is the parameter representing an assumption about the proportion of causal markers. For variant weight calculations, we used LDPred-Fast, the sparsified prediction method described by Mefford et al. [34]. For each trialed ρ value in the set = [0.001, 0.003, 0.01, 0.03, 0.1, 0.3, 1.0], PD-GPRS scores were first calculated for validation set subjects followed by prediction of PD status as a function of age, gender, and PD-GPRS was tested using a logistic regression model, with area under the curve (AUC) performance reported following fivefold cross-validation. The ρ parameter found to achieve the best AUC performance was ρ = 1.0; PD-GPRS scores for further analysis were thus computed for Test Set subjects under the assumption ρ = 1.0.

### QUANTIFICATION AND STATISTICAL ANALYSIS

Test set data not previously used to select ρ for PD-GPRS score calculation was used to evaluate PD-GPRS prediction of PD. We first performed an exploratory data analysis comparing PD-GPRS in Control versus PD Case subjects using two-sample t-test. To further investigate if GPRS predicts risk of PD, we then built a logistic regression model to calculate the odds ratio of being diagnosed with PD comparing high-percentile GPRS individuals with all subjects in percentiles below this cutoff, taking gender and age into account as confounding factors in the model. Cumulative incidence plots of age of onset versus prevalence of PD were also examined, separated as the top and bottom deciles of PD-GPRS scores and a grouping of the middle 2-9 deciles. PD Case and Control subjects in the Unified AMP-PD cohort also had available standardized motor function assessments: MDS-UPDRS parts 2 and 3. Mean scores for PD Case and Control cohorts were compared by permutation test for the top and bottom 10% of PD-GPRS, and mean Parts II and III scores by PD-GPRS decile were plotted to examine whether any trend between PD-GPRS and motor function might be observed.

#### Two-tail Analysis

Genetic attributes enriched for among samples in the two tails of PD-GPRS may provide valuable insights. Samples in the top 5% and bottom 5% of GPRS among Test set were considered as the high GPRS group and low GPRS group, respectively. Genetic attributes were then compared for high GPRS versus low GPRS subjects, with identification of those alleles which were observed with significantly greater odds among high-PD-GPRS individuals by Fisher’s exact test.

### PATHWAY ANALYSIS

Nearest genes to variants identified as occurring with significantly greater frequency among the top 5% of PD-GPRS score individuals were explored for pathway enrichment using the Enrichment Network analysis tool available in NetworkAnalyst [25].

### KEY RESOURCES TABLE

| **Deposited Data** |  |
| --- | --- |
| Accelerating Medicines Partnership:<br>Parkinson's Disease | https://amp-pd.org/ |
| FinnGen | https://www.finngen.fi/en |
| **Software and Algorithms** |  |
| Terra Platform | https://terra.bio/ |
| R/RStudio | https://www.r-project.org/<br>https://rstudio.com/ |
| Python | https://www.anaconda.com/ |
| LDPred | https://github.com/bvilhjal/ldpred |

## REFERENCES

1. Tysnes, O.B. and A. Storstein, Epidemiology of Parkinson’s disease. J Neural Transm (Vienna), 2017. 124(8): p. 901–905.

2. Blauwendraat, C., et al., Parkinson’s disease age at onset genome-wide association study: Defining heritability, genetic loci, and alpha-synuclein mechanisms. Mov Disord, 2019. 34(6): p. 866–875.

3. Dorsey, E.R., et al., The Emerging Evidence of the Parkinson Pandemic. J Parkinsons Dis, 2018. 8(1): p. S3–S8.

4. Rizzo, G., et al., Accuracy of clinical diagnosis of Parkinson disease: A systematic review and meta-analysis. Neurology, 2016. 86(6): p. 566–76.

5. Billingsley, K.J., et al., Genetic risk factors in Parkinson’s disease. Cell Tissue Res, 2018. 373(1): p. 9–20.

6. Do, C.B., et al., Web-based genome-wide association study identifies two novel loci and a substantial genetic component for Parkinson’s disease. PLoS Genet, 2011. 7(6): p. e1002141.

7. Singleton, A.B., et al., alpha-Synuclein locus triplication causes Parkinson’s disease. Science, 2003. 302(5646): p. 841.

8. Blauwendraat, C., M.A. Nalls, and A.B. Singleton, The genetic architecture of Parkinson’s disease. Lancet Neurol, 2020. 19(2): p. 170–178.

9. Gan-Or, Z., et al., Differential effects of severe vs mild GBA mutations on Parkinson disease. Neurology, 2015. 84(9): p. 880–7.

10. Paul, K.C., et al., Association of Polygenic Risk Score With Cognitive Decline and Motor Progression in Parkinson Disease. JAMA Neurol, 2018. 75(3): p. 360–366.

11. Pihlstrom, L., et al., A cumulative genetic risk score predicts progression in Parkinson’s disease. Mov Disord, 2016. 31(4): p. 487–90.

12. Escott-Price, V., et al., Polygenic risk of Parkinson disease is correlated with disease age at onset. Ann Neurol, 2015. 77(4): p. 582–91.

13. Li, W.W., et al., Association of the Polygenic Risk Score with the Incidence Risk of Parkinson’s Disease and Cerebrospinal Fluid alpha-Synuclein in a Chinese Cohort. Neurotox Res, 2019. 36(3): p. 515–522.

14. Ibanez, L., et al., Parkinson disease polygenic risk score is associated with Parkinson disease status and age at onset but not with alpha-synuclein cerebrospinal fluid levels. BMC Neurol, 2017. 17(1): p. 198.

15. Jacobs, B.M., et al., Parkinson’s Disease Determinants, Prediction and Gene-Environment Interactions in the UK Biobank 2020.

16. Nalls, M.A., et al., Identification of novel risk loci, causal insights, and heritable risk for Parkinson’s disease: a meta-analysis of genome-wide association studies. Lancet Neurol, 2019. 18(12): p. 1091–1102.

17. Khera, A.V., et al., Polygenic Prediction of Weight and Obesity Trajectories from Birth to Adulthood. Cell, 2019. 177(3): p. 587–596 e9.

18. Khera, A.V., et al., Genome-wide polygenic scores for common diseases identify individuals with risk equivalent to monogenic mutations. Nat Genet, 2018. 50(9): p. 1219–1224.

19. Vilhjalmsson, B.J., et al., Modeling Linkage Disequilibrium Increases Accuracy of Polygenic Risk Scores. Am J Hum Genet, 2015. 97(4): p. 576–92.

20. Choi, S.W., T.S.H. Mak, and P.F. O’Reilly, A Guide to Performing Polygenic Risk Score Analyses. 2018: https://www.biorxiv.org/.

21. AMP-PD, AMP-PD.

22. FinnGen, FinnGen Documentation of R4 release.

23. Siitonen, A., et al., Finnish Parkinson’s disease study integrating protein-protein interaction network data with exome sequencing analysis. Sci Rep, 2019. 9(1): p. 18865.

24. Goetz, C.G., et al., Movement Disorder Society-sponsored revision of the Unified Parkinson’s Disease Rating Scale (MDS-UPDRS): scale presentation and clinimetric testing results. Mov Disord, 2008. 23(15): p. 2129–70.

25. Zhou, G., et al., NetworkAnalyst 3.0: a visual analytics platform for comprehensive gene expression profiling and meta-analysis. Nucleic Acids Res, 2019. 47(W1): p. W234–W241.

26. Keller, M.F., et al., Using genome-wide complex trait analysis to quantify ‘missing heritability’ in Parkinson’s disease. Hum Mol Genet, 2012. 21(22): p. 4996–5009.

27. Brudek, T., Inflammatory Bowel Diseases and Parkinson’s Disease. J Parkinsons Dis, 2019. 9(2): p. S331–S344.

28. Li, X., J. Sundquist, and K. Sundquist, Subsequent risks of Parkinson disease in patients with autoimmune and related disorders: a nationwide epidemiological study from Sweden. Neurodegener Dis, 2012. 10(1-4): p. 277–84.

29. Witoelar, A., et al., Genome-wide Pleiotropy Between Parkinson Disease and Autoimmune Diseases. JAMA Neurol, 2017. 74(7): p. 780–792.

30. Bye, C.R., et al., Axonal Growth of Midbrain Dopamine Neurons is Modulated by the Cell Adhesion Molecule ALCAM Through Trans-Heterophilic Interactions with L1cam, Chl1, and Semaphorins. J Neurosci, 2019. 39(34): p. 6656–6667.

31. Lewis, C.M. and E. Vassos, Polygenic risk scores: from research tools to clinical instruments. Genome Med, 2020. 12(1): p. 44.

32. Ibanez, L., et al., Polygenic Risk Scores in Neurodegenerative Diseases: a Review. Current Genetic Medicine Reports, 2019. 7(1): p. 22–29.

33. Genomes Project, C., et al., A global reference for human genetic variation. Nature, 2015. 526(7571): p. 68–74.

34. Mefford, J., et al., Efficient Estimation and Applications of Cross-Validated Genetic Predictions to Polygenic Risk Scores and Linear Mixed Models. J Comput Biol, 2020. 27(4): p. 599–612.

